# A Two-Stage Multimodal Contrastive Framework for PET-Based Prediction of Obstructive Coronary Artery Disease

**DOI:** 10.64898/2026.08.20.26360938

**Authors:** Shiva Mostafavi, Aakash Shanbhag, Giselle Ramirez, Mark Lemley, Robert J. H. Miller, Panithaya Chareonthaitawee, Joanna X. Liang, Damini Dey, Paul B. Kavanagh, Leandro Slipczuk, Mark I. Travin, Erick Alexanderson, Isabel Carvajal Juarez, Rene R. S. Packard, Mouaz H. Al-Mallah, Andrew J. Einstein, Terrence D. Ruddy, Robert A. deKemp, Kevin Boczar, Attila Feher, Ronny R. Buechel, Wanda Acampa, Stacey Knight, Viet T. Le, Thomas L. Rosamond, Daniel S. Berman, Marcelo F. Di Carli, Piotr Slomka

**Author notes:** Co-senior authors (Marcelo F. Di Carli, Piotr Slomka). **Address for correspondence:** Piotr J. Slomka, PhD, FACC Cedars-Sinai Medical Center 8700 Beverly Blvd Los Angeles, CA 90048.

## Abstract

**Background:** Positron emission tomography (PET) myocardial perfusion imaging (MPI) provides complementary information on perfusion, myocardial blood flow and ventricular function. While these markers are often considered collectively during interpretation, their quantitative integration with imaging and clinical data into a unified predictive framework remains limited. We developed a multimodal artificial intelligence framework that combines PET polar maps with quantitative imaging and clinical features to improve obstructive coronary artery disease (CAD) detection.

**Methods:** We retrospectively analyzed the multicenter REFINE PET registry. Among 38,682 PET MPI studies from 14 sites, 2,833 patients without known prior CAD underwent invasive coronary angiography within 180 days. Obstructive CAD was defined as ≥50% left main stenosis or ≥70% stenosis in other major epicardial coronary arteries. We developed a two-stage contrastive learning framework to learn multimodal PET representations from studies without angiographic labels and transfer them to supervised CAD prediction. In Stage 1, PET image and tabular encoders were pretrained on 12,225 PET MPI studies from eight development sites using 15-channel PET polar maps, quantitative PET perfusion, flow and gated functional measures, and clinical variables. In Stage 2, the pretrained encoders and a lightweight classification head were fine-tuned in 968 angiography-labeled patients, using lower encoder learning rates to limit overfitting. The model was externally validated for angiographically defined obstructive CAD detection in 1,865 patients from six independent sites and compared with standard PET MPI metrics.

**Results:** The prevalence of obstructive CAD was 60% in the training cohort (66% male, median age of 70 years [63, 77]), and 55% in the external validation cohort (64% male, median age of 67 years [60–74]). In external validation, the AI model achieved an AUC of 0.85 (95% confidence interval (CI), 0.83–0.87) for obstructive CAD detection and outperformed conventional quantitative PET metrics (all P < 0.001). At a specificity matched to visual summed stress score, the AI model achieved higher sensitivity (89% [95% CI, 87–91] versus 85% [95% CI, 82–87]) and negative predictive value (81% [95% CI, 77–84] versus 73% [95% CI, 69–77]; both p<0.001). The overall net reclassification improvement was 8.9% (95% CI, 4.2–13.6%; p = 0.001).

**Conclusions:** Multimodal contrastive pretraining improved obstructive CAD detection from PET imaging beyond conventional perfusion-based scoring in independent multisite external validation.

**Key question:** Can multimodal contrastive pretraining leverage PET MPI studies without angiographic labels to improve detection of angiography-defined obstructive CAD by integrating PET polar maps, quantitative PET/CT measures, and clinical variables?

**Key results:** The AI model achieved the highest external AUC for obstructive CAD detection, outperforming SSS and other PET-derived metrics. At an SSS-matched threshold, it improved sensitivity, negative predictive value, reclassification, and net benefit.

**Take-home message:** Multimodal contrastive pretraining leverages PET studies without angiographic labels and improves obstructive CAD detection by integrating polar maps with quantitative PET/CT and clinical data.

**Graphical abstract:** 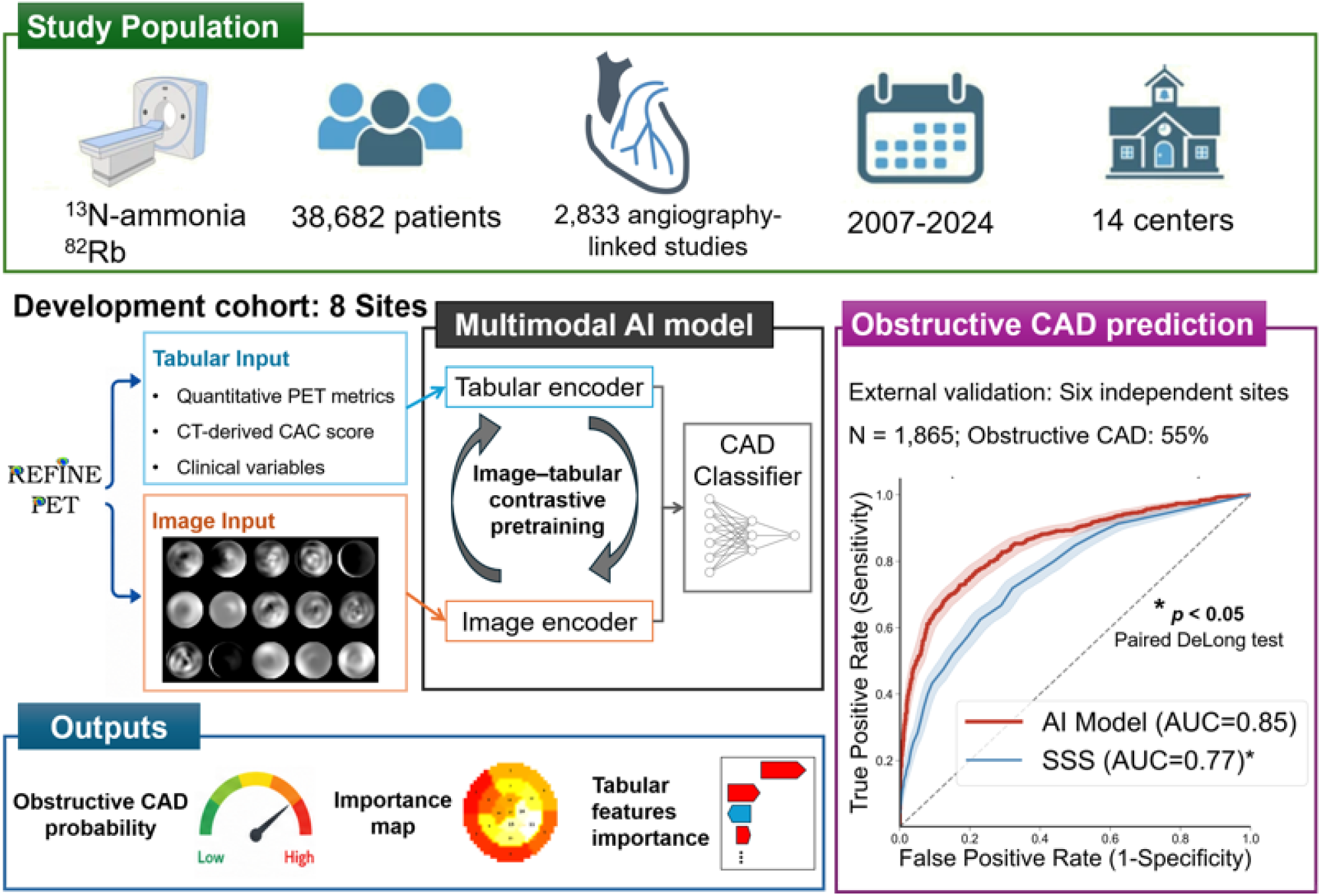

A contrastively pretrained multimodal AI model integrated PET polar maps, quantitative PET metrics, CT-derived CAC, and clinical variables to predict obstructive CAD, improve performance over SSS, and provide patient-level attribution maps. SSS indicates summed stress score.

## Introduction

Coronary artery disease (CAD) remains a leading cause of morbidity and mortality worldwide[1]. Invasive coronary angiography (ICA) provides the anatomical basis for assessing coronary stenosis [2], however its invasive nature limits its use for broad diagnostic evaluation. Accurate non-invasive identification of obstructive CAD is therefore essential for selecting patients who may benefit from further invasive testing and treatment [3].

Positron emission tomography myocardial perfusion imaging (PET MPI) enables comprehensive non-invasive CAD assessment by providing complementary measures of coronary and myocardial physiology within a single examination, including regional relative perfusion, rest and stress myocardial blood flow (MBF), myocardial flow reserve (MFR), left ventricular function, and coronary artery calcium burden when computed tomography attenuation correction (CTAC) is available. Accurate diagnosis of obstructive CAD requires consideration of these physiological, anatomical, and clinical data[4]. While relative perfusion is a cornerstone technique for diagnostic assessment, it may underestimate multivessel disease[5]. Reduced MFR may reflect obstructive epicardial stenosis, diffuse atherosclerosis, coronary microvascular dysfunction, or a combination of these processes[6]. The presence of calcium, while indicative of atherosclerosis, is not an accurate predictor of obstructive CAD. Physicians interpreting PET MPI often need to integrate physiological, anatomical, and clinical findings that may provide conflicting signals, making assessment of obstructive CAD challenging.

Artificial intelligence (AI) may help resolve this diagnostic challenge by integrating the physiological, anatomical, and clinical information available from PET MPI into a unified predictive framework. While previous deep-learning models have been developed for automated detection CAD from perfusion imaging[7–12], they relied solely on supervised labels derived from invasive angiography. However, such labels are available for only a small proportion of patients undergoing PET MPI, leaving many clinically acquired studies unlabeled and therefore unavailable for conventional supervised model training.

Contrastive learning provides a way to learn from these data without endpoint-specific labels. It trains representations by aligning matched observations and separating mismatched observations in a shared embedding space. Contrastive language–image pretraining (CLIP)[13] extended this principle across modalities by aligning images with their corresponding text descriptions. More recently, multimodal pretraining of volumetric CT with radiology reports and electronic health record data demonstrated that routinely paired clinical information can support transferable representations before task-specific fine-tuning[14].

In this study, we developed a two-stage multimodal framework that aligns PET MPI polar maps with quantitative imaging-derived and clinical features through contrastive pretraining, without requiring angiographic CAD labels. The pretrained encoders were then utilized for the downstream task of predicting angiographically defined obstructive CAD. We hypothesized that this approach would learn transferable representations of MPI and improve diagnostic performance compared with conventional PET-derived markers and an otherwise identical supervised model trained from random initialization. We also assessed patient-level image and tabular feature attribution to characterize the clinical information learned by the model.

## Methods

### Study population

In this multicenter retrospective study, we included patients undergoing cardiac PET MPI from the REgistry of Flow and Perfusion Imaging for Artificial Intelligence with positron emission tomography (REFINE PET)[15] between 2007 and 2024. The full registry included 38,682 PET MPI studies from 14 clinical sites. Of these, 37,438 studies had both rest and stress PET images available and were eligible for cohort assignment. Sites were divided a priori into model-development and independent external-validation groups to ensure site-level separation between training and testing.

Eight sites contributed to the internal development cohort, which included 14,357 PET MPI studies. For Stage 1 contrastive pretraining, 12,225 studies with available clinical data and CT attenuation-correction data were used; angiographic CAD labels were not used during this stage. Within the development sites, 968 patients without known prior CAD who underwent invasive coronary angiography within 180 days of PET MPI formed the ICA-labeled subset used for supervised CAD fine-tuning in Stage 2.

Six independent sites were reserved for external validation and contributed 23,081 PET MPI studies, with no patient or site overlap with the development cohort. Among these, 1,865 patients without known prior CAD who underwent invasive coronary angiography within 180 days of PET MPI formed the external diagnostic validation cohort. Obstructive CAD was defined as ≥50% stenosis of the left main coronary artery or ≥70% stenosis in other epicardial coronary arteries on invasive angiography.

Institutional review boards (IRB) approval was obtained at each site, and the study complies with the Declaration of Helsinki. Sites either obtained written informed consent or waiver of consent for the use of the de-identified data.

#### PET/CT Protocol

All patients underwent same-day rest and pharmacologic stress PET myocardial perfusion imaging using ^82^Rb or ^13^N-ammonia. Studies were acquired using a Biograph mCT 64 PET/CT scanner (Siemens Healthineers, Erlangen, Germany), Discovery RX (GE Healthcare, Waukesha, Wisconsin, USA), Discovery MI (GE Healthcare, Waukesha, Wisconsin, USA), Discovery STE (GE Healthcare, Waukesha, Wisconsin, USA), Ingenuity TF (Philips Healthcare, Best, the Netherlands), or GEMINI TF (Philips Healthcare, Best, the Netherlands). A 6-minute rest list-mode acquisition was started immediately before administering weight-based doses of ^82^Rb or ^13^N-ammonia. Concurrent with the beginning of the injection, a 6-minute stress imaging acquisition was initiated. Prior to each PET acquisition for rest and stress, a low-dose helical CT scan was performed for attenuation correction[16]. Patients were scanned in the supine position and images were reconstructed using vendor-recommended reconstruction algorithms optimized for each scanner and site. CT images were acquired with site-specific imaging protocols[15].

#### PET image quantification

All PET imaging variables, including myocardial perfusion, blood flow, ejection fraction, and transient ischemic dilation (TID) ratio, were computed automatically in batch mode at the core laboratory with dedicated software (QPET, Cedars-Sinai Medical Center, Los Angeles, CA) [15]. Rest and stress relative perfusion were quantified using total perfusion deficit (TPD), and normal myocardial perfusion was defined as stress TPD <5%[17]. Rest and stress MBF were measured using a 1-tissue compartment kinetic model for ^82^Rb PET and a 2-compartment model for ^13^N-ammonia PET. MBF and the spillover fraction from the blood to the myocardium were determined via numeric optimization [18–20]. Stress and rest flow values, expressed in units of mL/g/min, were computed for each sample on the polar map. Minimal segmental stress MBF (Stress MBF) was used in the AI model. Integrated MFR (iMFR) was derived by jointly evaluating regional relative perfusion and absolute MFR, classifying myocardium into normal, focally impaired, focally preserved, or diffusely impaired[21]. The percentage of focally impaired myocardium was evaluated as a comparator for obstructive CAD detection.

#### CTAC image analysis

CAC was quantified from CTAC scans using our previously validated deep learning framework [22, 23], followed by calculation of CAC scores according to established methodology[24]. The model generated a total patient-level CAC score by identifying calcified lesions within the cardiac region and summing the lesion-level scores across all analyzed slices. The resulting total CAC score was log-transformed and used as a continuous measure of coronary calcification burden in subsequent analyses.

#### Clinical scoring

PET/CT scans were visually assessed during clinical reporting by experienced physicians at each site with access to available clinical and imaging data. Perfusion abnormalities were summarized using summed stress scores (SSS), summed rest scores (SRS), and summed difference score (SDS) using the 17-segment American Heart Association model[25]. SSS was used as the primary clinical perfusion comparator because of its standardized role in summarizing stress perfusion abnormality.

### Model inputs and preprocessing

The model used paired image and tabular inputs from each PET MPI study. Image inputs consisted of a 15-channel PET polar-map stack incorporating relative perfusion, absolute myocardial blood flow, myocardial flow reserve, spillover, wall motion, wall thickening, and phase and amplitude features derived from ECG-gated acquisitions. Tabular inputs included quantitative PET/CT variables and clinical variables collected at the time of PET MPI. Quantitative PET/CT variables included stress and rest perfusion measures, stress and rest MBF, MFR, TID, gated ventricular-function measures, and log-transformed CAC score. Clinical variables included age, sex, and body mass index (BMI). The final analytic cohorts included only studies with complete image and tabular inputs required by the model.

### Model architecture and training strategy

A two-stage multimodal deep-learning framework was developed to detect angiography-defined obstructive CAD from cardiac PET MPI. The framework first learned transferable patient-level representations without using angiographic CAD labels and subsequently adapted these representations for supervised CAD prediction. Image and tabular encoders transformed the PET polar maps and corresponding tabular variables into patient-level embeddings. Detailed encoder architectures and implementation parameters are provided in the Supplementary Methods.

#### Stage 1, multimodal contrastive pretraining

The image and tabular encoders were trained jointly using a symmetric contrastive learning objective adapted from the CLIP framework[13]. Within each training batch, pairwise cosine similarities were computed between all PET polar map embeddings and all corresponding clinical and quantitative feature embeddings, forming an image–tabular similarity matrix. Same-patient image–tabular combinations defined the target matches for the contrastive loss, whereas cross-patient combinations served as nonmatching comparisons within the loss function. Similarities were scaled by a learnable temperature parameter, and cross-entropy loss was computed in both image-to-clinical and clinical-to-image directions. This objective encouraged the model to align PET imaging patterns with corresponding patient-level physiological and clinical profiles without requiring angiographic CAD labels. The resulting pretrained encoders were subsequently transferred to the supervised classification stage (Figure 1).

**Figure 1.**
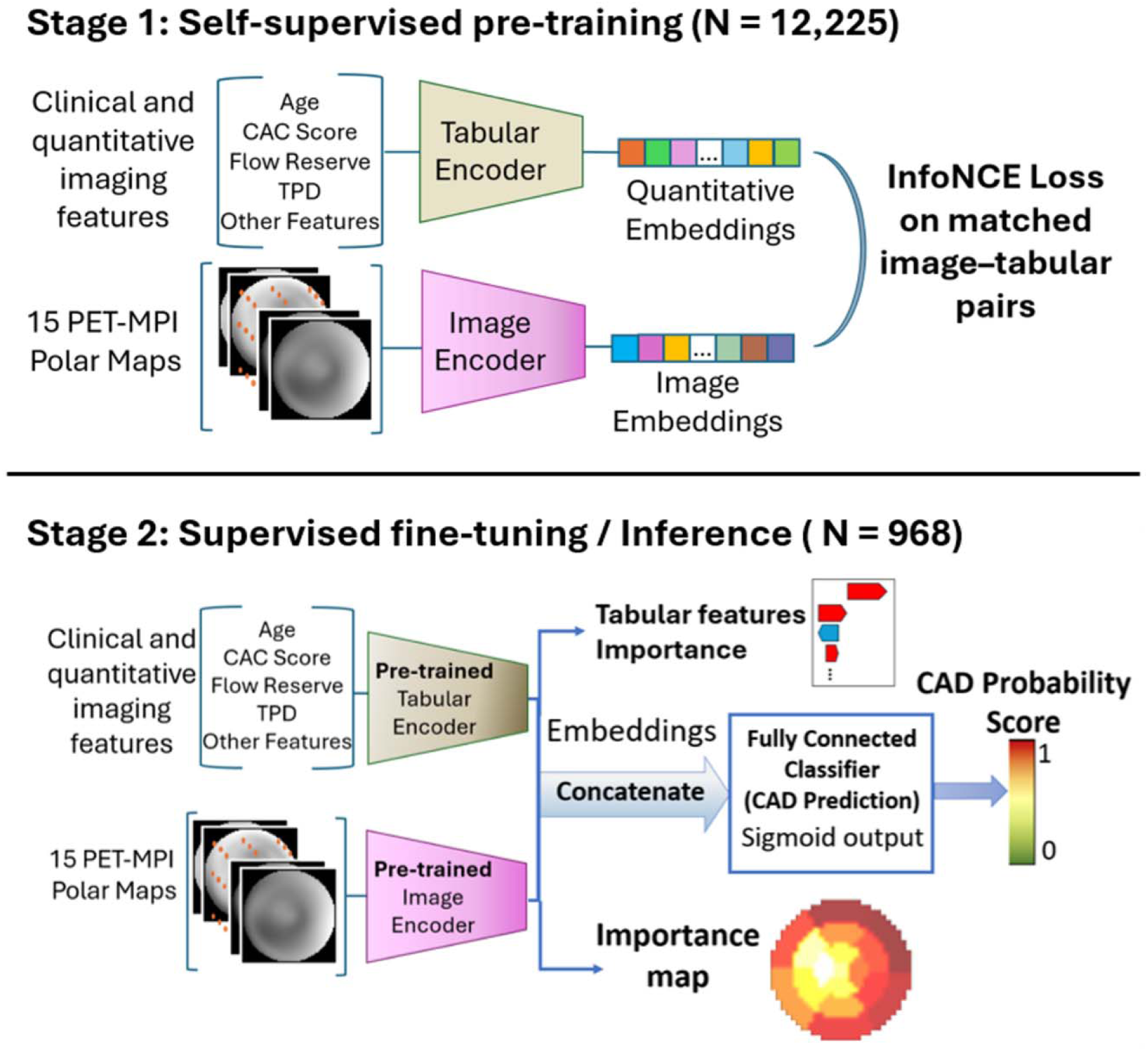
Overview of the proposed two-stage multimodal contrastive learning framework for obstructive coronary artery disease (CAD) prediction. Stage 1: self-supervised pre-training: Clinical and quantitative imaging features and 15-channel PET MPI polar maps were encoded using separate tabular and image encoders. A symmetric contrastive objective based on InfoNCE loss aligned matched image–tabular representations from the same patient, while cross-patient image–tabular combinations served as nonmatching comparisons within the loss function, enabling label-free multimodal representation learning. **Stage 2: supervised fine-tuning and inference**. The pretrained image and tabular embeddings were concatenated and passed to a fully connected classifier, with the output logit converted by a sigmoid function into the predicted probability of obstructive CAD. During inference, integrated gradients and SHAP were used to provide patient-level attribution of image regions and tabular features, respectively. CAD, coronary artery disease; CAC, coronary artery calcium; InfoNCE, information noise-contrastive estimation; MPI, myocardial perfusion imaging; SHAP, Shapley additive explanations; TPD, total perfusion deficit.

#### Stage 2, supervised CAD classification

The pretrained image and tabular encoders were adapted for angiography-defined obstructive CAD prediction. Their outputs were concatenated and passed through a fully connected classification head to produce a continuous CAD logit. The pretrained encoders were fine-tuned using a lower learning rate than the newly initialized classification head to allow gradual task-specific adaptation while limiting disruption of representations learned during contrastive pretraining. The model was trained using binary cross-entropy with logits loss, and the output logit was transformed using a sigmoid function into an AI-CAD probability ranging from 0 to 1, with higher values indicating a greater predicted likelihood of obstructive CAD.

### Evaluation

The trained models were evaluated in a separate external validation cohort from six independent clinical sites, with no patient, site, or temporal overlap with the development cohort.

#### Frozen-embedding evaluation

To assess clinically relevant information encoded in the pretrained representation, the image encoder was frozen and evaluated using linear-probe and nearest-neighbor analyses for angiography-defined CAD and five clinical outcomes: heart failure, major adverse cardiovascular events, all-cause mortality, cardiovascular mortality, and myocardial infarction. Detailed methods are provided in the Supplementary Methods.

#### Fine-tuned model evaluation

For supervised evaluation, the fine-tuned model generated a continuous AI-CAD probability for each patient in the external diagnostic validation cohort. Discrimination for angiography-defined obstructive CAD was assessed using the area under the receiver operating characteristic curve and compared with conventional PET-derived metrics. For threshold-dependent evaluation, the AI-CAD operating threshold was selected in the 10% internal validation set to match the specificity of SSS ≥4 [26], then fixed and applied unchanged to the external validation cohort, as detailed in the Supplementary Methods. All threshold-dependent metrics, including sensitivity, specificity, predictive values, and reclassification measures, were calculated at this prespecified operating point.

#### Model interpretation

To interpret model decision-making and move beyond a black-box prediction, we applied two complementary attribution methods to the fine-tuned model: integrated gradients, which attribute the predicted CAD probability to input polar map pixels by integrating gradients[27] along a path from a baseline input to the actual input, and kernel SHAP[28], which estimates each input feature’s marginal contribution to the prediction using a weighted linear approximation of the model’s local behavior. Both methods were applied to the 15-channel polar map representation to localize the myocardial regions and importance of the clinical features to evaluate whether image- and tabular-derived predictions were supported by clinically plausible regional and quantitative information.

#### Ablation analyses and model robustness

Ablation analyses were performed to evaluate the contribution of contrastive pretraining. The proposed AI model was first compared with an otherwise identical model trained from scratch, with Kaiming initialization for convolutional layers and Xavier initialization for linear layers, using the same supervised training procedure and external validation cohort. The proposed model was compared with a previously developed AI model based only on PET-derived quantitative features, without imaging polar maps[7] and evaluated in the same external cohort.

Model robustness was evaluated across the six external clinical sites by calculating site-specific AUCs. Prespecified subgroup analyses were also performed according to PET radiotracer (^82^Rb versus ^13^N-ammonia), sex (men versus women), and BMI category (<30 versus ≥30 kg/m²). Within each subgroup, the AUC of the AI model was compared with SSS.

To assess label efficiency, patient-level subsets comprising 10%, 25%, 50%, and 100% of the fine-tuning cohort were sampled with stratification by site and obstructive CAD status. At each fraction, identical model architectures were trained using either contrastively pretrained or random initialization, with matched patient subsets and fixed validation and external cohorts. The pretraining dataset was unchanged, and models were evaluated in same external cohort.

### Statistical analysis

Continuous variables were summarized as median with interquartile range (IQR; Q1–Q3), and categorical variables were reported as counts and percentages. Baseline characteristics were compared between groups using the Wilcoxon rank-sum test for non-normally distributed continuous variables and Pearson’s χ² test for categorical variables.

Model discrimination was quantified using the area under the receiver operating characteristic curve (AUC-ROC) with 95% confidence intervals, and AUCs were compared between models using the pairwise DeLong test[29]. Threshold-dependent performance metrics, including sensitivity, specificity, positive predictive value, and negative predictive value, were calculated at the validation-derived AI-CAD operating threshold described above. Paired differences in threshold-dependent metrics were compared using McNemar’s test. The incremental classification value of the AI-CAD score relative to SSS was assessed using the net reclassification improvement (NRI). The event and non-event components of the NRI were calculated separately and summed to obtain the overall NRI. Percentile confidence intervals were estimated using stratified bootstrap resampling, and two-sided empirical bootstrap *P* values were calculated as twice the smaller proportion of bootstrap estimates falling on either side of zero.

Decision curve analysis was performed in the external validation cohort to evaluate the clinical utility of the AI model across threshold probabilities for obstructive CAD. Net benefit was calculated across thresholds from 0.01 to 0.90 and compared with SSS ≥4, treat-all, and treat-none strategies. In a post hoc exploratory analysis, each conventional PET metric was evaluated at its predefined abnormal threshold, and an AI-CAD threshold was selected within the external cohort to match that metric’s observed specificity. Performance was then compared at each matched-specificity operating point. Calibration of predicted probabilities was assessed using calibration curves and the Brier score. Prespecified subgroup analyses were performed as described above. Confidence intervals for all performance metrics were estimated via bootstrapping, and p value <0.05 was considered statistically significant. All statistical analyses were performed with Pandas (version 2.1.1), Numpy (version 1.24.3), Scipy (version 1.11.4), Lifelines (version 0.28.0) and Scikit-learn (version 1.3.0) in Python 3.12.5 (Python Software Foundation, Wilmington, DE, USA).

## Results

### Study population

Baseline demographic and clinical characteristics are summarized in Table 1. The pretraining cohort included 12,225 PET studies. Of these, 968 patients who underwent ICA within 180 days of PET myocardial perfusion imaging were included in stage 2. The fine-tuning dataset comprised patients who were 66% male, with a median age of 70 [63, 77] years; 61% had obstructive CAD. The external validation cohort included 1,865 patients from six independent sites that were not used during training. This cohort was 64% male, had a median age of 67 [60, 74] years, and 55% had obstructive CAD.

**Table 1:** Patient characteristics of the pretraining, supervised training, and external validation cohorts. Continuous variables are presented as median (interquartile range), and categorical variables as number (percentage). P values compare the supervised training cohort with the external validation cohort.

| Variable | Pretrain set | Train (fine tuning) | External Cohort | p-value |
| --- | --- | --- | --- | --- |
| N | 12,225 | 968 | 1,865 |  |
| Age | 69 (61, 77) | 70 (63, 77) | 67 (60, 74) | <0.001 |
| BMI, kg/m<sup>2</sup> | 27.80 (24.60, 32.28) | 27.94 (24.59, 32.30) | 31.00 (26.54, 37.00) | <0.001 |
| **Sex** |  |  |  | 0.369 |
| Female | 4479 (36.6%) | 330 (34.1%) | 669 (35.9%) |  |
| Male | 7746 (63.4%) | 638 (65.9%) | 1196 (64.1%) |  |
| Smoking | 1664 (13.6%) | 145 (15.0%) | 559 (30.0%) | <0.001 |
| Hypertension | 9023 (73.8%) | 727 (75.1%) | 1526 (82.4%) | <0.001 |
| Diabetes Mellitus | 4378 (35.8%) | 400 (41.3%) | 757 (40.7%) | 0.335 |
| Dyslipidemia | 8536 (69.8%) | 652 (67.4%) | 1349 (72.8%) | <0.001 |
| Family History | 1849 (15.1%) | 153 (15.8%) | 630 (34.3%) | <0.001 |
| Peripheral vascular disease | 1703 (13.9%) | 90 (9.3%) | 321 (17.5%) | <0.001 |
| **Race** |  |  |  | <0.001 |
| American Indian or Alaska Native | 35 (0.3%) | 4 (0.4%) | 15 (0.8%) |  |
| Asian | 462 (3.8%) | 35 (3.6%) | 30 (1.6%) |  |
| Black or African American | 1887 (15.4%) | 138 (14.3%) | 130 (6.7%) |  |
| Native Hawaiian or<br>Other Pacific Islander | 43 (0.4%) | 4 (0.4%) | 20 (1.1%) |  |
| White | 8639 (70.7%) | 711 (73.1%) | 1578 (84.6%) |  |
| Unavailable | 1159 (9.5%) | 76 (7.9%) | 92 (4.9%) |  |
| **Isotope** |  |  |  | 0.936 |
| <sup>13</sup>N-ammonia | 2991 (24.5%) | 297 (30.7%) | 568 (30.5%) |  |
| <sup>82</sup>Rb | 9234 (75.5%) | 671 (69.3%) | 1297 (69.5%) |  |
| **Pharmacological Stress**<br>**Agent** |  |  |  | <0.001 |
| Dipyridamole | 860 (7.0%) | 54 (5.6%) | 168 (9.0%) |  |
| Adenosine | 1615 (13.2%) | 165 (17.0%) | 83 (4.5%) |  |
| Regadenoson | 9717 (79.5%) | 747 (77.2%) | 1580 (84.7%) |  |
| Dobutamine | 32 (0.3%) | 2 (0.2%) | 34 (1.8%) |  |
| Missing | 1 (0.0) | 0 (0.0) | 0 (0.0) |  |
| **PET-CT quantitative image analysis parameters** |  |  |  |  |
| Stress TPD | 3.63 (1.22, 9.74) | 8.41 (3.81, 16.27) | 9.35 (4.61, 17.38) | 0.027 |
| Ischemic TPD | 2.61 (0.98, 5.75) | 5.98 (2.87, 11.11) | 6.17 (3.19, 11.71) | 0.129 |
| TID | 1.04 (0.97, 1.11) | 1.06 (0.99, 1.15) | 1.05 (0.99, 1.13) | 0.035 |
| Stress MBF | 1.57 (1.09, 2.05) | 1.27 (0.82, 1.72) | 1.17 (0.81, 1.61) | <0.001 |
| Myocardial Flow reserve | 1.76 (1.30, 2.24) | 1.44 (1.01, 1.94) | 1.48 (1.08, 1.96) | 0.115 |
| Coronary calcium score | 269.19 (16.13,<br>1246.76) | 387.19 (54.68, 1358.91) | 219.68 (12.22, 843.50) | <0.001 |
| End Diastolic Volume | 99.87 (77.78, 130.51) | 107.54 (85.78, 142.05) | 113.55 (84.93, 149.89) | 0.021 |
| End Systolic Volume | 32.17 (19.59, 53.28) | 39.11 (24.81, 67.54) | 41.30 (25.56, 71.02) | 0.301 |
| Stress LVEF, % | 68 (57, 76) | 64 (49, 71) | 63 (49, 73) | 0.75 |
| Rest LVEF, % | 65 (54, 72) | 62 (50, 70) | 62 (48, 71) | 0.858 |
| **Outcome** |  |  |  |  |
| Obstructive CAD | - | 586 (60.5%) | 1021 (54.7%) | 0.004 |

BMI, body mass index; TPD, total perfusion deficit; TID, transient ischemic dilation; MBF, myocardial blood flow; MFR, myocardial flow reserve; LVEF, left ventricular ejection fraction; CAD, coronary artery disease.

### Evaluation of pretrained image representations

Using frozen pretrained image embeddings, both the similarity-weighted 50-nearest-neighbor analysis and the linear probe achieved an AUC of 0.81 (95% CI, 0.79–0.83) and significantly outperformed SSS (P<0.001). Performance of the frozen representations across additional clinical outcomes is reported in Supplementary Table 1.

### Fine-tuned model

The hybrid AI model achieved the highest overall discrimination for obstructive CAD detection, with an AUC of 0.85, and significantly outperformed all prespecified PET-derived comparators, including SSS, ischemic TPD, stress TPD, flow measures, Integrated MFR, and CAC score (all P < 0.001; Figure 2).

**Figure 2:**
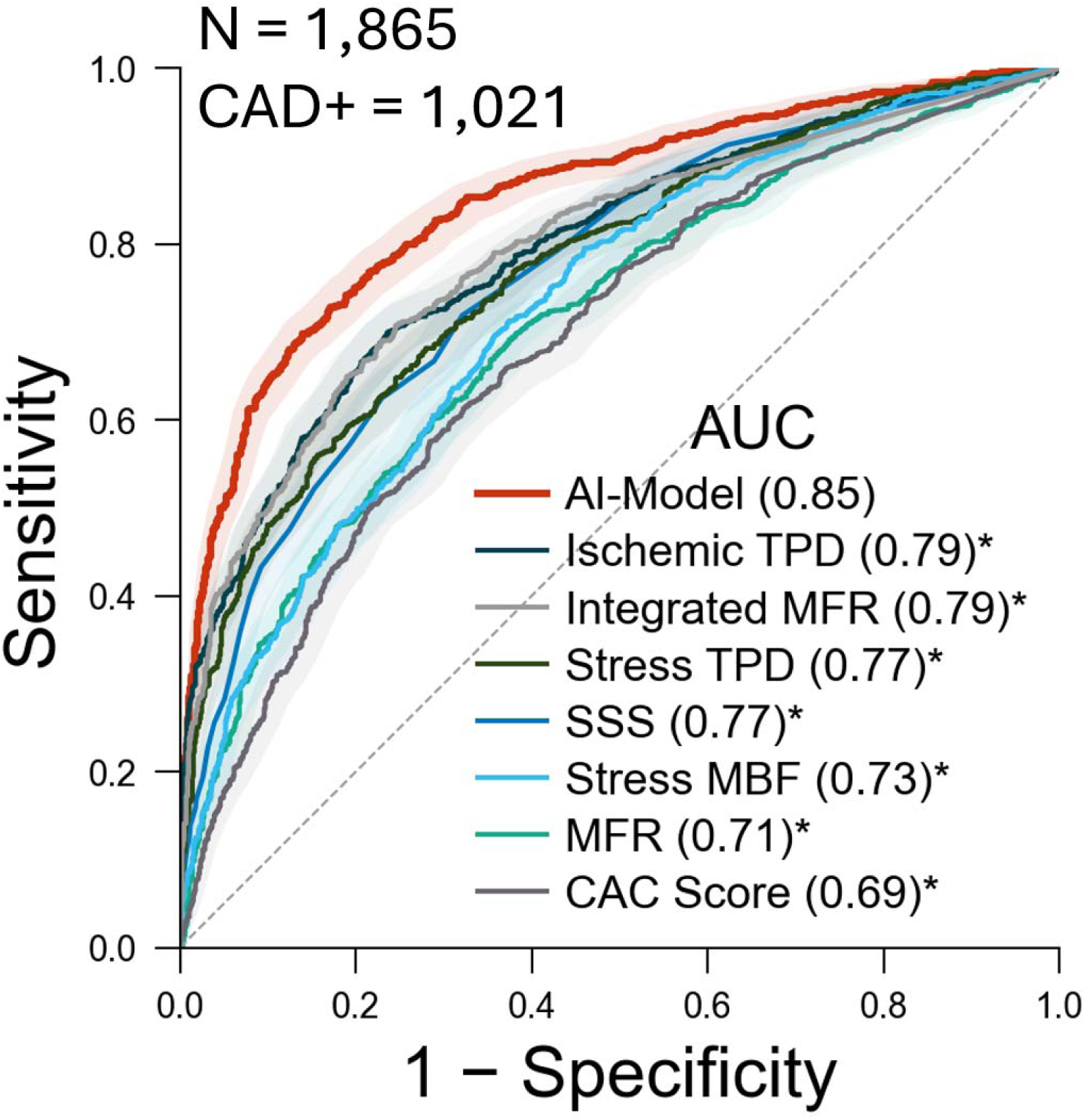
Diagnostic performance of the AI model. Receiver operating characteristic curves compare the AI model with conventional PET-derived metrics for obstructive CAD detection. Shaded regions indicate 95% confidence intervals. **\* Asterisks** indicate a significant difference in AUC compared with the AI model by DeLong’s test (P < 0.001). Integrated MFR represents the extent of focally impaired myocardium, and MFR and MBF were selected from the minimum of the 3 vessels. AUC, area under the receiver operating characteristic curve; CAC, coronary artery calcium; MBF, myocardial blood flow; MFR, myocardial flow reserve; SSS, summed stress score; TPD, total perfusion deficit.

The AI-CAD operating threshold was selected in the internal validation set to match the specificity of SSS ≥4 and was then applied unchanged to the external validation cohort. At this prespecified operating point, the AI model achieved higher sensitivity than SSS (89% versus 85%; P < 0.001) and higher negative predictive value (0.81 versus 0.73; P <0.001; Figure 3A). At the SSS-matched operating point, the AI model also demonstrated the highest sensitivity and negative predictive value among the conventional PET-derived metrics evaluated at their predefined operating thresholds (all P < 0.05) (Table 2). Patient-level reclassification showed improvement in both CAD-positive and CAD-negative patients. The AI model showed an event NRI of 4.4% and a non-event NRI of 4.5%, resulting in an overall NRI of 8.9% (P = 0.001; Figure 3B–C). Decision curve analysis showed that the AI model provided greater net benefit than SSS ≥4 across the evaluated threshold range and exceeded treat-all and treat-none strategies across most clinically relevant threshold probabilities (Supplementary Figure 2).

**Figure 3:**
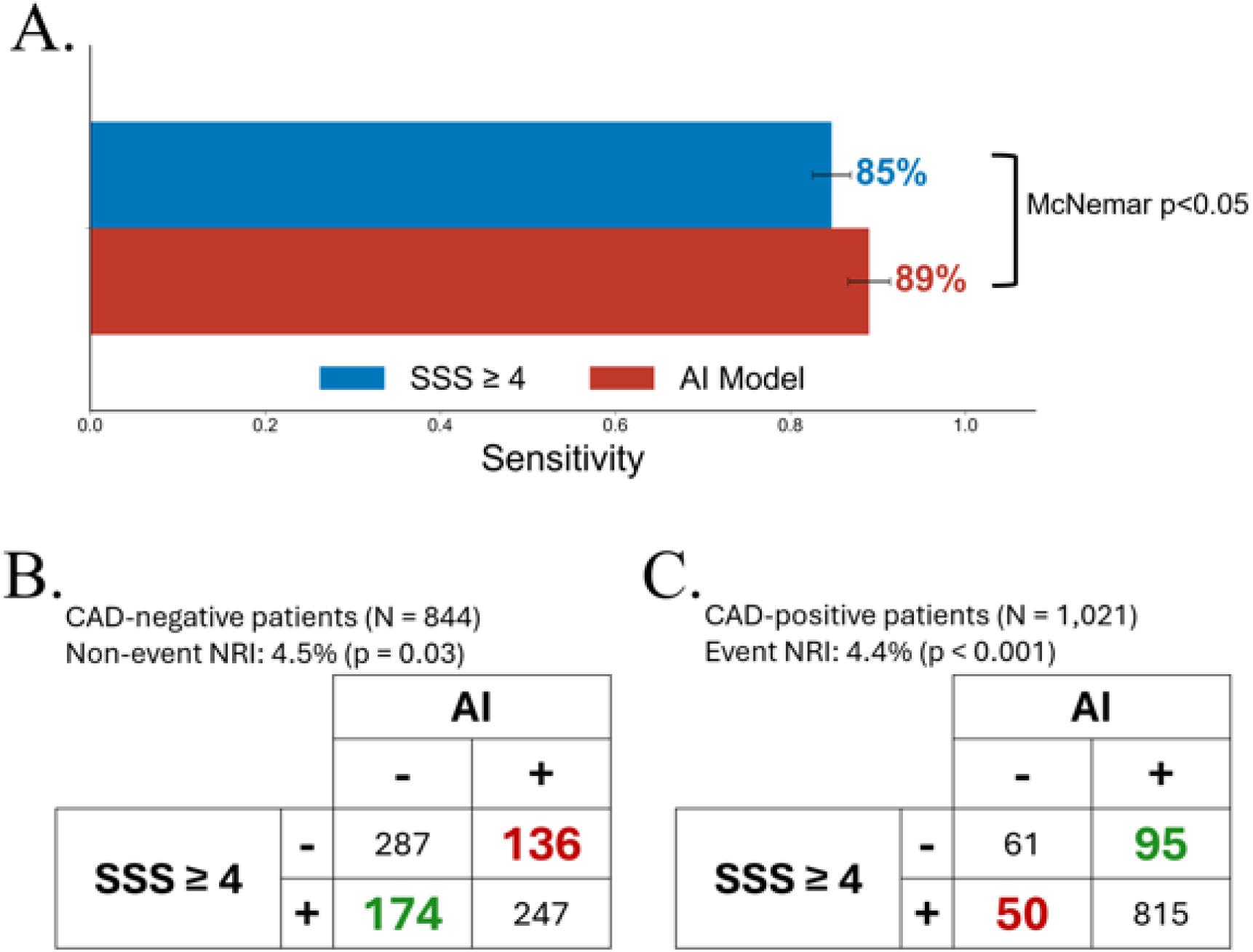
Sensitivity and reclassification of the AI model compared with SSS. A) Sensitivity of the AI model and SSS ≥4 for detecting obstructive CAD in the external validation cohort. The AI-CAD threshold was selected in the internal validation set to match the specificity of SSS ≥4 and then applied unchanged to the external validation cohort. Error bars indicate 95% confidence intervals; the paired difference in sensitivity was assessed using McNemar’s test. B–C) Reclassification of CAD-negative and CAD-positive patients by the AI model compared with SSS ≥4. Non-event NRI was 4.5% (P = 0.03), event NRI was 4.4% (P < 0.001), and overall NRI was 8.9%. CAD, coronary artery disease; NRI, net reclassification improvement; SSS, summed stress score.

**Table 2:** Diagnostic performance at the SSS-matched operating point. The AI-CAD threshold was selected in the internal validation set to match the specificity of the conventional criterion SSS ≥4, then fixed and applied unchanged to the external validation cohort. Asterisks indicate significant differences by McNemar’s test (P < 0.05).

| **Model** | **Sensitivity**<br>**(95% CI)** | **Specificity**<br>**(95% CI)** | **PPV**<br>**(95% CI)** | **NPV**<br>**(95% CI)** |
| --- | --- | --- | --- | --- |
| AI model | 89 (87-91) | 55 (51-58) | 70 (68-73) | 81 (77-84) |
| Ischemic TPD * | 78 (75-80) | 62 (59-65) | 71 (69-74) | 70 (67-73) |
| Stress TPD * | 86 (84-88) | 45 (41-48) | 65 (63-68) | 72 (68-76) |
| MFR* | 73 (70-76) | 55 (51-58) | 66 (63-69) | 63 (59-66) |
| Stress MBF* | 74 (72-77) | 58 (55-61) | 68 (65-71) | 65 (62-68) |
| SSS * | 85 (82-87) | 50 (47-53) | 67 (65-70) | 73 (69-77) |

CAD, coronary artery disease; CI, confidence interval; NPV, negative predictive value; PET, positron emission tomography; PPV, positive predictive value. MFR and MBF were selected from the minimum of the 3 vessels. MBF, myocardial blood flow; MFR, myocardial flow reserve; TPD, total perfusion deficit; SSS, summed stress score.

At specificity matched separately to each conventional PET metric, the AI model consistently achieved higher sensitivity, indicating that it identified more patients with obstructive CAD at the same false-positive burden. The differences in sensitivity were significant across all comparisons (all P < 0.05) (Supplementary Figure 3).

In the external cohort, the prevalence of CAD increased steadily across AI-score quintiles, from 16.9% in the lowest-risk group to 95.4% in the highest-risk group (Figure 4). Calibration-in-the-large was preserved, with a calibration intercept of 0, but the calibration slope was 1.83 and the Brier score was 0.166, indicating imperfect probability calibration with overestimation in lower-risk groups and underestimation in higher-risk groups.

**Figure 4:**
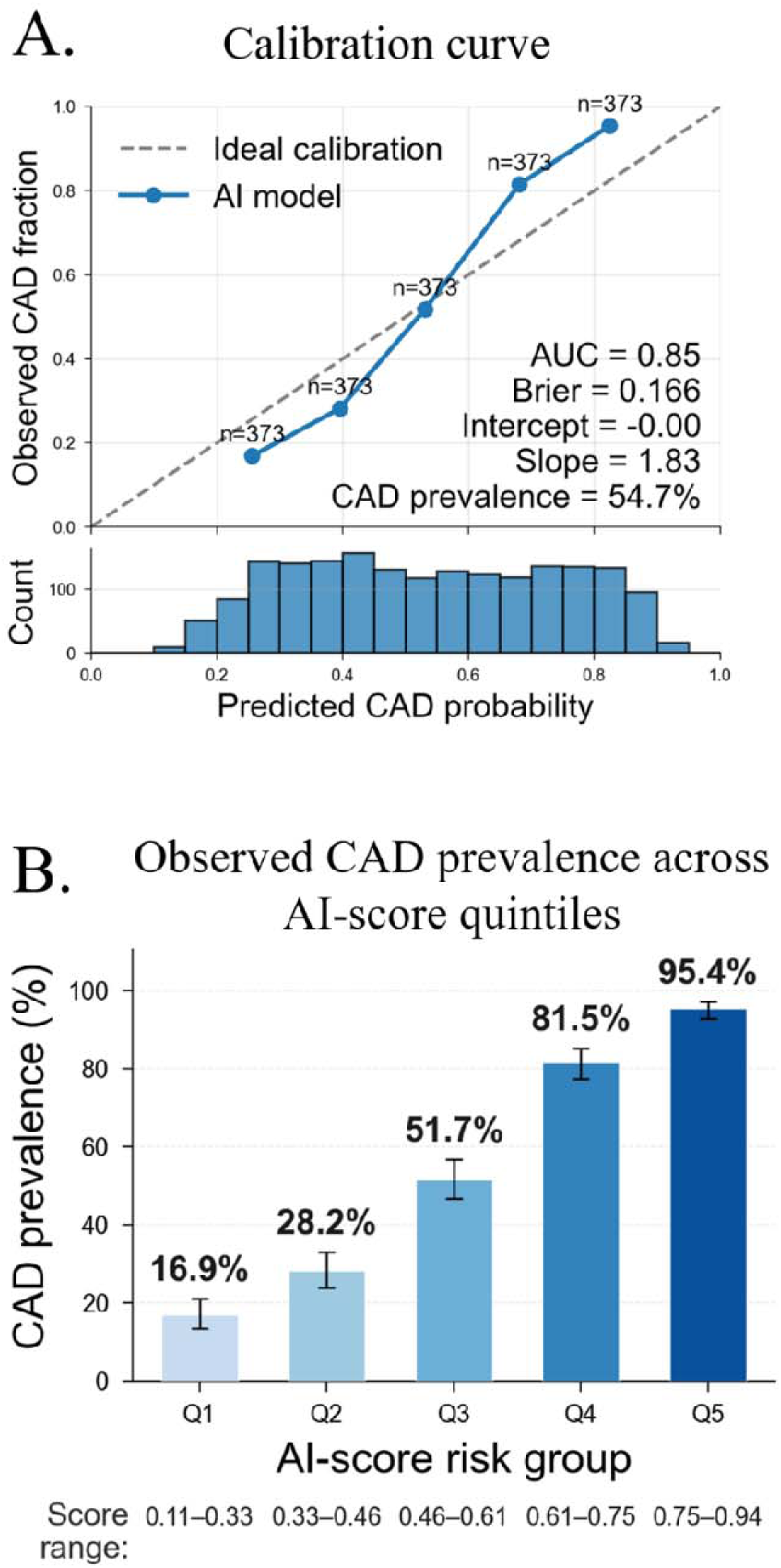
Calibration and risk stratification of the AI model in the external cohort. A) **Calibration curve** and distribution of predicted CAD probabilities in the external validation cohort. B) **Observed CAD prevalence across AI-score quintiles**. Error bars indicate 95% confidence intervals.

### Interpretability and clinical plausibility: patient-level model interpretation

To illustrate patient-level model interpretability, a representative CAD-positive case (Figure 5) that was missed by clinical score (SSS = 0) was shown. The patient was a female in her 70’s with BMI of 40 kg/m² and obstructive CAD on angiography, including 60% left main stenosis and 85% LAD stenosis. The AI model assigned a CAD probability above the diagnostic threshold, correctly identifying the patient as CAD-positive. Post hoc image attribution using integrated gradients demonstrated increased importance in the LAD territory, consistent with the angiographic finding of severe LAD stenosis. SHAP-based tabular feature attribution showed that MFR was the largest tabular contributor pushing the prediction toward CAD, followed by CAC score.

**Figure 5:**
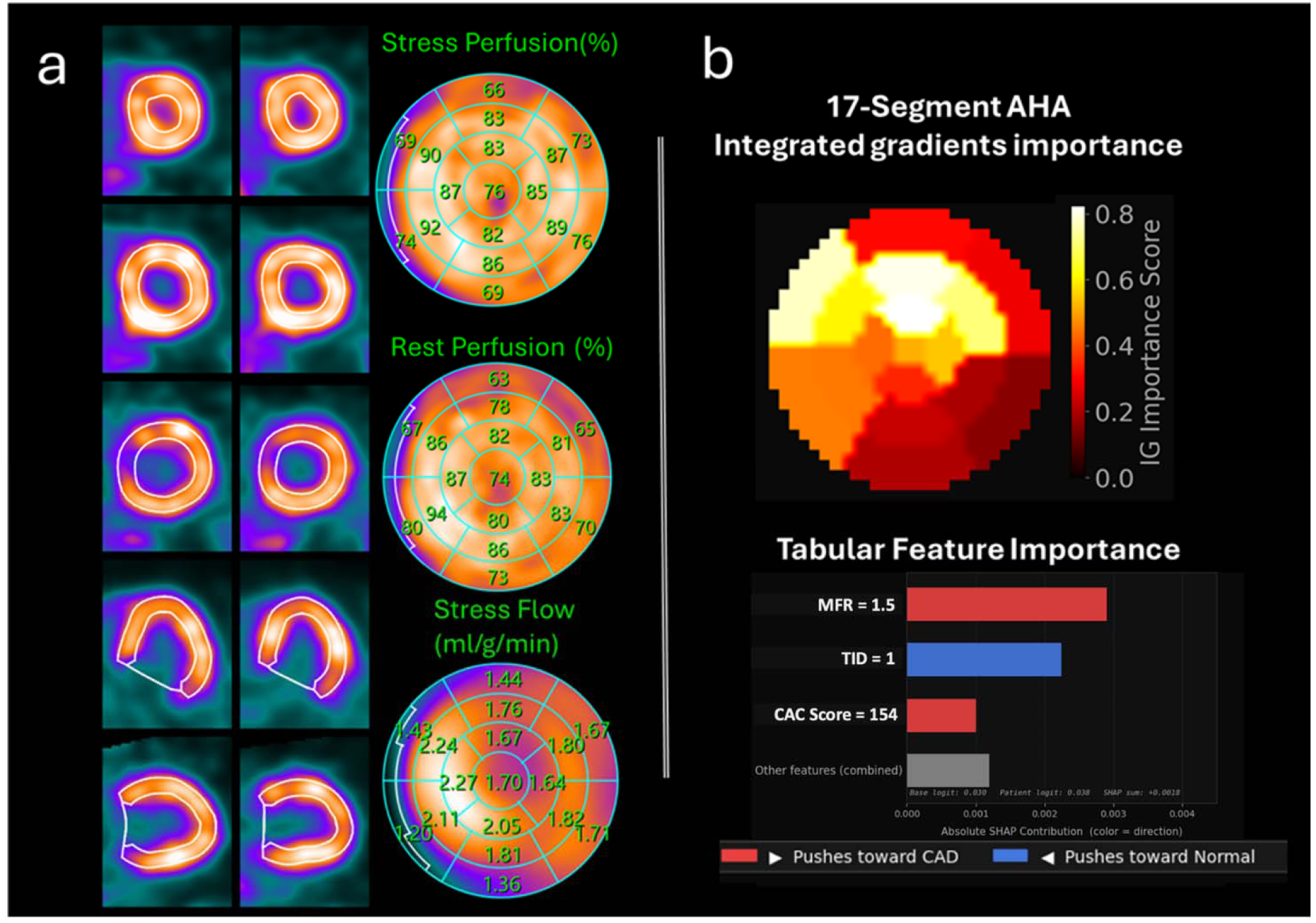
CAD-positive case: Female in her 70’s (BMI 40 kg/m²), with severe stenosis on angiography **(LM: 60%, LAD 85%)**. Despite a clinically normal perfusion score (SSS = 0), the AI model predicted a **high CAD probability**, correctly reflecting the true disease status. (Myocardial flow reserve (MFR)= 1.5, stress total perfusion deficit (TPD) = 3%, and CAC score = 154.) a) Representative PET MPI images and polar maps showing stress perfusion, rest perfusion, and stress myocardial blood flow. b) Integrated gradients highlighted the LAD territory, consistent with angiographic disease, while SHAP-based tabular attribution identified reduced MFR and CAC score as contributors toward CAD prediction; TID contributed toward a normal classification. AHA, American Heart Association; CAC, coronary artery calcium; LAD, left anterior descending artery; MFR, myocardial flow reserve; TID, transient ischemic dilation.

### Ablation analyses and model robustness

Contrastive pretraining significantly improved obstructive CAD discrimination compared with an otherwise identical model trained from random initialization (AUC, 0.85 [95% CI, 0.83–0.87] versus 0.83 [95% CI, 0.81–0.85]; P=0.03). The proposed model also significantly outperformed a previous AI model for CAD detection based on PET-derived quantitative features, which achieved an AUC of 0.83 (95% CI, 0.81–0.85) in the same external cohort (P = 0.02)[7].

The AI model maintained higher discrimination than SSS across external sites and prespecified subgroups (Figure 6). Site-specific AUCs ranged from 0.76 to 0.90 for the AI model, and the AI model outperformed SSS at five of the six sites (all P < 0.05); at the remaining site the difference did not reach statistical significance (0.80 versus 0.74; P = 0.071). Discrimination was also higher across radiotracer subgroups, with AUCs of 0.86 for ^82^Rb and 0.82 for ^13^N-ammonia, compared with 0.78 and 0.73 for SSS, respectively. Similar findings were observed in men and women and across BMI categories (<30 and ≥30 kg/m²), with the AI model outperforming SSS in all subgroup comparisons (all P < 0.001). Detailed subgroup-specific AUCs, 95% confidence intervals, and sample sizes are provided in Supplementary Table 2.

**Figure 6:**
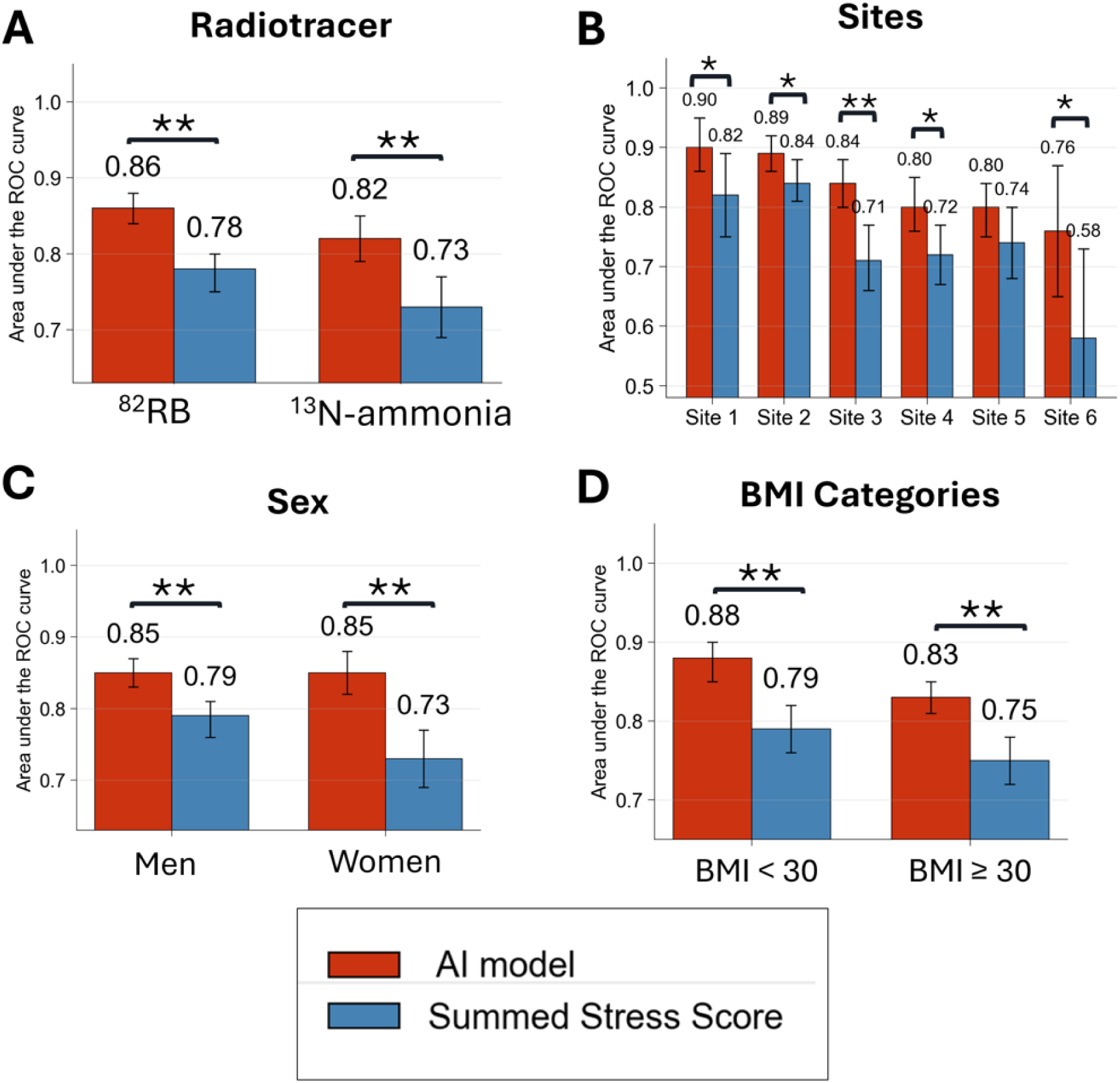
Subgroup diagnostic performance of the AI model compared with SSS. AUCs are shown for the AI model and summed stress score across subgroups defined by A, radiotracer (^82^Rb and ^13^N-ammonia); B, external validation site; C, sex; and D, BMI category (<30 and ≥30 kg/m²). Error bars indicate 95% confidence intervals. Asterisks indicate significant differences in AUC between the AI model and SSS by paired DeLong testing (* P < 0.05; ** P < 0.001). AUC, area under the receiver operating characteristic curve; BMI, body mass index; SSS, summed stress score.

The contrastively pretrained model maintained stable external discrimination across labeled-data fractions, with AUCs ranging from 0.83 to 0.85, whereas the randomly initialized model improved progressively from 0.74 with 10% of the fine-tuning cohort to 0.83 with the complete cohort (Supplementary Figure 4).

## Discussion

In this multicenter study, we developed and externally validated a multimodal contrastive learning framework that integrates spatial PET polar maps with quantitative imaging and clinical data while leveraging abundant PET studies without angiographic labels. In the external diagnostic cohort of 1,865 patients, including 1,021 with angiography-confirmed obstructive CAD, the AI model achieved an AUC of 0.85 and outperformed established PET-derived markers, including SSS, perfusion-defect measures, MFR, stress MBF, and CAC score.

Performance was consistent across sites, isotopes, sex, and body mass index. These findings indicate that jointly learning from spatial and quantitative PET information captures clinically relevant disease patterns that are not fully represented by individual imaging markers.

The improvement over individual imaging metrics supports the value of integrated assessment and is consistent with prior studies showing that combining multiparametric MPI information improves diagnostic and prognostic performance[7, 8, 21, 30–38]. Zhang et al[7] developed an XGBoost model integrating perfusion, flow, and CAC measurements for obstructive CAD detection and outperforming individual PET biomarkers. Similarly, a prior ^15^O-water PET/CT study showed that an explainable deep-learning model integrating polar maps with clinical, CTA, and PET variables achieved performance comparable to clinical reading for flow-limiting CAD[39].

Related evidence from SPECT MPI also supports the value of spatial polar-map information for AI-based CAD detection. Machine-learning approaches combining SPECT MPI polar maps with clinical data have achieved diagnostic performance comparable to expert interpretation[35].

Using the multicenter REFINE-SPECT registry[36], Otaki et al. integrated perfusion, functional, and clinical features, outperforming standard SPECT metrics and clinical interpretation[8]. Betancur et al. further showed that deep learning models using raw and quantitative perfusion polar maps improved per-patient and per-vessel CAD detection compared with TPD[40].

Another SPECT polar-map AI study also reported improved AUC for CAD detection[41]. Together, these studies support the value of multimodal AI analysis in MPI.

Extending these approaches, our model combined quantitative PET-derived features with spatial polar-map information. It outperformed both the iMFR-derived percentage of focally impaired myocardium[21], a composite measure integrating regional MFR and ischemic TPD, and a previously established AI model for obstructive CAD detection based on PET-derived biomarkers in the same external cohort[7], supporting the added value of spatial imaging information beyond tabular measurements alone.

A major limitation of conventional supervised AI development is its dependence on large datasets with high-quality disease labels. This is particularly a limitation for angiography-confirmed CAD because only a selected subset of patients undergoing PET MPI undergo invasive coronary angiography. Consequently, most routinely acquired PET studies cannot be used directly for supervised CAD model training. Contrastive pretraining addressed this limitation by treating each patient’s polar maps and quantitative profile as a matched pair, allowing the image encoder to learn representations related to perfusion, myocardial blood flow, ventricular function, calcium burden, and clinical characteristics without requiring angiographic labels. Similar multimodal approaches have shown that combining cardiac magnetic resonance images with clinical data improves downstream cardiovascular diagnosis[42], and multimodal pretraining of volumetric CT with radiology reports and electronic health record codes enabled zero-shot and transferable performance across multiple clinical tasks. Together, these studies demonstrate that routinely paired clinical modalities can provide effective supervision when endpoint-specific labels are limited[14].

Our study extends contrastive representation learning to PET MPI and provides evidence that the pretrained model captured clinically meaningful patterns beyond those learned through conventional supervised training alone. The improvement over random initialization suggests that exposure to large-scale multimodal PET data established a more informative starting representation for obstructive CAD detection. Consistent with this, the pretrained model maintained stable external AUCs of 0.83–0.85 when fine-tuned with 10%–100% of the labeled cohort, highlighting the potential of pretraining when labeled data is limited. In addition, the performance of a linear probe applied to frozen image embeddings indicates that CAD-related information was already organized within the pretrained latent space before task-specific encoder adaptation.

The improvement in discrimination also translated into better patient-level classification. Compared with clinical SSS, the AI model correctly reclassified 95 patients with obstructive CAD upward and 174 patients without obstructive CAD downward. This bidirectional reclassification suggests that the model identified additional CAD-positive patients missed by SSS while also correcting some false-positive classifications. However, the effect on downstream testing or treatment cannot be determined from the selected operating point alone. Future studies should evaluate prespecified rule-in and rule-out thresholds, and their negative predictive value, false-negative rate, and impact on clinical decision-making.

Limited interpretability remains a major barrier to the clinical adoption of AI. We therefore incorporated complementary image- and tabular-attribution methods into the inference workflow to provide patient-level explanations. Integrated gradients localized regions of the PET polar maps that contributed to individual predictions, and SHAP-based analysis estimated the direction and magnitude of contributions from quantitative and clinical features, which can help clinicians assess whether an AI prediction is supported by physiologically plausible information and may facilitate review of conflicting cases.

Several limitations define the next steps toward clinical translation. First, the retrospective design introduces selection and referral bias because fewer than 10% of registry patients underwent invasive angiography and were included in supervised diagnostic evaluation. Prospective studies in broader referral populations are therefore required. Second, the reference standard was based on anatomical stenosis and did not include fractional flow reserve; future work should incorporate functional coronary indices when available and evaluate related phenotypes such as microvascular dysfunction and diffuse non-obstructive disease. Third, the interval between PET MPI and angiography may have allowed changes in disease status or treatment. Calibration was imperfect in the external diagnostic cohort, with a Brier score of 0.166; however, because this cohort consisted of patients selected for invasive angiography, calibration should be reassessed in broader PET MPI populations with different disease prevalence and risk distributions.

Similarly, positive and negative predictive values may not be directly transportable to unselected PET MPI populations with lower disease prevalence, whereas AUC, sensitivity, and specificity are less dependent on disease prevalence. In addition, pairwise comparisons across imaging metrics, sites, and subgroups were not adjusted for multiplicity and should be interpreted as exploratory. Per-vessel analysis was not performed because the primary model was developed for patient-level obstructive CAD classification, and several model inputs and comparators, including SSS, LVEF, TID, and CAC score were summarized at the patient level. Future work should evaluate territory-specific or vessel-level model outputs against LAD, LCX, and RCA angiographic disease. Although most processing was automated, myocardial contour quality control was performed by an experienced technologist; future studies should evaluate fully automated processing and sensitivity to contouring errors. The prognostic analyses were observational and cannot establish that AI-guided management improves outcomes. Finally, training on a single GPU demonstrated computational feasibility but limited evaluation of larger architectures and broader scaling strategies.

### Conclusions

In summary, PET MPI contains more clinically relevant information than is captured by individual metrics or by the small subset of studies with angiographic labels. Multimodal contrastive pretraining provides a practical way to use that information. By learning from routinely acquired PET studies and then adapting to an angiographic endpoint, the proposed framework improved obstructive CAD detection and generalized across independent sites. These findings support data-efficient multimodal representation learning as a promising direction for comprehensive PET MPI interpretation.

## Disclosures

PJS, DD, and DB declare equity interest in APQ Health and participates in software royalties for QPS software at Cedars-Sinai Medical Center. PJS has also received research grant support from Siemens Medical Systems and consulting fees from Synektik S.A. DB participates in software royalties for QPS software at Cedars-Sinai Medical Center. DB has also received research grant support from the Dr. Miriam and Sheldon G. Adelson Medical Research Foundation and served as a consultant for GE Healthcare. MDC received consulting fees from MedTrace, Valo Health and IBA and institutional grant support from Sun Pharma, Xylocor and Intellia. RM received consulting fees from Alnylam and Bayer and research support from Alberta Innovates. PC received consulting fees from GE Healthcare, Cardiovascular Clinical Sciences, and IBA, and royalties from UpToDate. AJE reports consulting for Artrya, authorship fees from Wolters Kluwer Healthcare—UpToDate, and serving on scientific advisory boards for Axcellant and Canon Medical Systems USA; his institution has grants/grants pending from Alexion, Attralus, BridgeBio, Canon Medical Systems USA, GE HealthCare, Intellia Therapeutics, International Atomic Energy Agency, Ionis Pharmaceuticals, National Institutes of Health, Pfizer, and Shockwave Medical. RRB has received speaker fees from GE Healthcare and Gilead and research grant support from GE Healthcare and the Swiss Heart Foundation. RRSP has received an investigator-initiated research grant from and serves as a consultant for GE HealthCare. RdK receives royalties from Rubidium PET technologies licensed to Jubilant Radiopharma and INVIA Medical Solutions; and received unrestricted research grant funding or honoraria from Siemens Molecular Imaging, IONETIX and Jubilant Radiopharma. LS received institutional grants from Amgen and Philips, honoraria from Elsevier for Editor-in-Chief position at Progress in Cardiovascular Diseases and reports equity in APQ Health Inc. VTL has received research grant support from J&J/Janssen, has received honorarium from the American College of Cardiology for Editor-in-Chief role at Cardiosmart, and has served on advisory boards for Amgen, Amarin, Bayer, Boehringer Ingelheim, Esperion, Idorsia, iRhythm, Merck, Novartis, Novonordisk, and Pfizer. The remaining authors have declared no competing interests.

## Funding

This research was supported in part by grant R35HL161195 from the National Heart, Lung, and Blood Institute/National Institutes of Health (NHLBI/NIH) and R01EB034586 from the National Institute of Biomedical Imaging and Bioengineering (PI: Piotr Slomka). The content is solely the responsibility of the authors and does not necessarily represent the official views of the National Institutes of Health.

## Data Availability

To the extent allowed by data sharing agreements and IRB protocols, the deidentified data and data analysis code from this manuscript will be shared upon written request

## List of abbreviations

AUC: area under the receiver operating characteristic curve
CAC: coronary artery calcium
CAD: coronary artery disease
CTAC: computed tomography attenuation correction
ICA: invasive coronary angiography
MBF: myocardial blood flow
MFR: myocardial flow reserve
PET MPI: positron emission tomography myocardial perfusion imaging
SSS: summed stress score
TPD: total perfusion deficit

## Notes

### Author Declarations

Institutional review boards (IRB) approval was obtained at each site, and the study complies with the Declaration of Helsinki. Sites either obtained written informed consent or waiver of consent for the use of the de-identified data. The investigators ensured that the institutional ethics committee at each center evaluated and approved the study protocol before data collection and transfer. The overall registries were approved by the Institutional Review Board (IRB) at Cedars-Sinai Medical Center (Office of Research Compliance and Quality Improvement).

