## Supplemental Material for "A Two-Stage Multimodal Contrastive Framework for PET-Based Prediction of Obstructive Coronary Artery Disease"

### Supplementary Methods

**Image encoder:** The image encoder was a convolutional neural network designed to extract spatial and multimodal information from the 15-channel polar map stack. The network included channel attention, a depthwise-separable convolutional stem, residual convolutional blocks, spatial attention, and dual global pooling. Channel attention was used to adaptively weight the contribution of each polar map modality before spatial feature extraction. The depthwise-separable stem first processed each input channel independently and then combined channel-wise information through pointwise convolution. This design preserved modality-specific spatial patterns before cross-channel integration. Three residual convolutional stages then extracted progressively higher-level regional and global myocardial features. A spatial attention module further emphasized informative myocardial regions within the final feature map. Finally, adaptive average pooling and adaptive max pooling were concatenated to generate a 1,024-dimensional image representation, capturing both global disease burden and focal high-intensity abnormalities. Then, this representation was passed through a nonlinear projection head to produce a 256-dimensional L2-normalized embedding.

**Tabular encoder:** Patient-level clinical and quantitative imaging features were encoded using a multilayer fully connected network. The encoder mapped the input feature vector to a 256-dimensional normalized embedding using sequential linear layers with batch normalization, ReLU activation, and dropout.

**Implementation:** Models were implemented in PyTorch and trained on a single NVIDIA TITAN RTX GPU with 24 GB of memory. Convolutional layers were initialized using Kaiming initialization, and linear layers were initialized using Xavier initialization. Batch normalization layers were initialized with unit scale and zero bias. Each polar-map channel was standardized independently using the mean and standard deviation calculated from the training cohort, and the same training-derived parameters were applied to the test cohorts. Model training used the Adam optimizer with weight decay, early stopping based on internal test AUROC, and separate training schedules for contrastive pretraining and supervised fine-tuning.

**Frozen-embedding evaluation of downstream tasks**

**Linear-probe analysis:** L2-normalized image embeddings were standardized using parameters estimated from the development training subset. For each endpoint, an L2-regularized logistic regression classifier was fitted to the frozen embeddings and corresponding binary outcome labels. No encoder parameters were updated or hyperparameters optimized. Performance was evaluated on the independent external cohort.

**Nearest-neighbor analysis:** L2-normalized embeddings were compared using cosine similarity, and the 50 most similar reference patients were identified; k=50 was selected using the internal development cohort and fixed before external evaluation. Each neighbor was weighted according to inverse cosine distance. For each endpoint e, a similarity-weighted score was calculated as

$$S_{q,e}= \frac{\sum_{i=1}^{50} w_{qi}y_{i,e}}{\sum_{i=1}^{50} w_{qi}}$$

where S_q,e_ is the similarity-weighted score for evaluation patient q and endpoint e, w_qi_ is the inverse cosine-distance weight assigned to reference patient i based on its similarity to patient q, and y_i,e_ is the binary outcome of reference patient i for endpoint e, coded as 1 when the endpoint was present and 0 otherwise.

**Threshold selection for AI-CAD classification**

The AI-CAD operating threshold was selected in the 10% internal validation set to match the observed specificity of SSS ≥4. The internal validation set included 97 patients, of whom 59 had obstructive CAD and 38 did not. The observed specificity of SSS ≥4 was 55.3% (95% CI, 39.7–69.9), yielding an AI-CAD probability threshold of 0.40. This threshold was fixed before evaluation and applied unchanged to the external validation cohort.

### Supplementary Tables

**Supplementary Table 1:** Prognostic discrimination of imaging-derived and frozen-embedding models for clinical outcomes. Area under the receiver operating characteristic curve (AUC) with 95% confidence intervals is reported for summed stress score (SSS), ischemic total perfusion deficit (iTPD), myocardial flow reserve (MFR), a linear probe trained on frozen image embeddings, and a k-nearest neighbors (KNN) classifier applied to the frozen embeddings. N denotes the number of patients included for each outcome, and Events denotes the number of observed events. Asterisks indicate significant differences in AUC between the quantitative metrics and linear probe by paired DeLong testing (P < 0.001).

| **Outcome** | **N**  **(Events)** | **SSS AUC (95% CI)** | **iTPD AUC (95% CI)** | **MFR AUC (95% CI)** | **Linear probe AUC (95% CI)** | **KNN AUC (95% CI)** |
| --- | --- | --- | --- | --- | --- | --- |
| MACE | 21,602  (6,457) | 0.63 *  (0.63–0.64) | 0.66 *  (0.65–0.66) | 0.68 *  (0.67–0.69) | 0.74  (0.73–0.75) | 0.73  (0.72–0.73) |
| All-cause death | 21,602  (4,409) | 0.60 *  (0.59–0.61) | 0.62 *  (0.61–0.63) | 0.70 *  (0.69–0.71) | 0.76  (0.76–0.77) | 0.74  (0.73–0.75) |
| Cardiovascular death | 18,590  (1,264) | 0.66 *  (0.64–0.68) | 0.68 *  (0.66–0.69) | 0.70 *  (0.68–0.71) | 0.78  (0.76–0.79) | 0.75  (0.73–0.76) |
| Myocardial infarction | 21,602  (1,415) | 0.67  (0.66–0.69) | 0.66  (0.65–0.68) | 0.64 *  (0.63–0.66) | 0.67  (0.66–0.68) | 0.67  (0.66–0.69) |
| Heart failure | 21,602  (1,884) | 0.65 *  (0.64–0.67) | 0.63 *  (0.62–0.64) | 0.67 *  (0.66–0.68) | 0.76  (0.75–0.77) | 0.73  (0.72–0.75) |

**Supplementary Table 2**. Subgroup diagnostic performance of the AI model compared with SSS. AUCs are shown with 95% confidence intervals. P values compare AUCs between the AI model and SSS within each subgroup using paired DeLong testing. AI, artificial intelligence; AUC, area under the receiver operating characteristic curve; BMI, body mass index; CAD, coronary artery disease; CI, confidence interval; SSS, summed stress score.

| **Subgroup** | **AI AUC (95% CI)** | **SSS AUC (95% CI)** | **Sample size (CAD+ %)** | **P value** |
| --- | --- | --- | --- | --- |
| **Site** |  |  |  |  |
| Site 1 | 0.90 (0.86–0.95) | 0.82 (0.75–0.89) | 159 (69%) | 0.014 |
| Site 2 | 0.89 (0.86–0.92) | 0.84 (0.81–0.88) | 597 (42%) | 0.004 |
| Site 3 | 0.84 (0.80–0.88) | 0.71 (0.66–0.77) | 324 (52%) | <0.001 |
| Site 4 | 0.80 (0.76–0.85) | 0.72 (0.67–0.77) | 438 (66%) | <0.001 |
| Site 5 | 0.80 (0.75–0.84) | 0.74 (0.68–0.80) | 283 (61%) | 0.071 |
| Site 6 | 0.76 (0.65–0.87) | 0.58 (0.43–0.73) | 64 (48%) | 0.011 |
| **Radiotracer** |  |  |  |  |
| ^82^Rb | 0.86 (0.84–0.88) | 0.78 (0.75–0.80) | 1,297 (52%) | <0.001 |
| ^13^N-ammonia | 0.82 (0.79–0.85) | 0.73 (0.69–0.77) | 568 (61%) | <0.001 |
| **Sex** |  |  |  |  |
| Men | 0.85 (0.83–0.87) | 0.79 (0.76–0.81) | 1,196 (60%) | <0.001 |
| Women | 0.85 (0.82–0.88) | 0.73 (0.69–0.77) | 669 (46%) | <0.001 |
| **BMI category** |  |  |  |  |
| BMI <30 kg/m² | 0.88 (0.85–0.90) | 0.79 (0.76–0.82) | 819 (57%) | <0.001 |
| BMI ≥30 kg/m² | 0.83 (0.81–0.85) | 0.75 (0.72–0.78) | 1,046 (53%) | <0.001 |

### Supplementary Figures

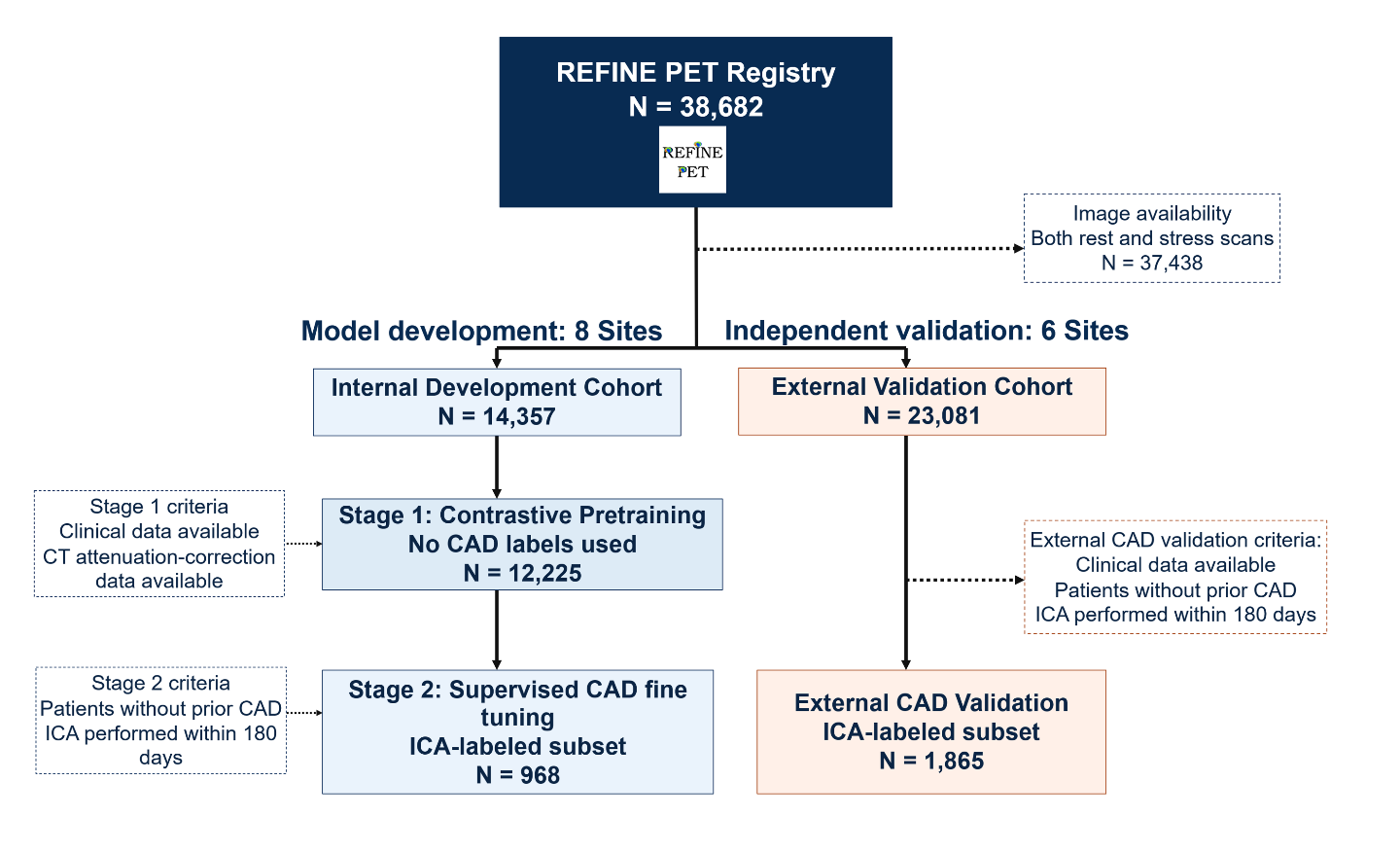
 **Supplementary Figure 1: Study design and cohort allocation.** The stage 1 of training included 12,225 PET MPI studies used for contrastive pretraining without CAD labels, with an invasive coronary angiography (ICA) subset of 968 patients used for supervised CAD classifier development. The external validation cohort included 23,081 patients from six independent sites, including an ICA subset of 1,865 patients used for external diagnostic validation.

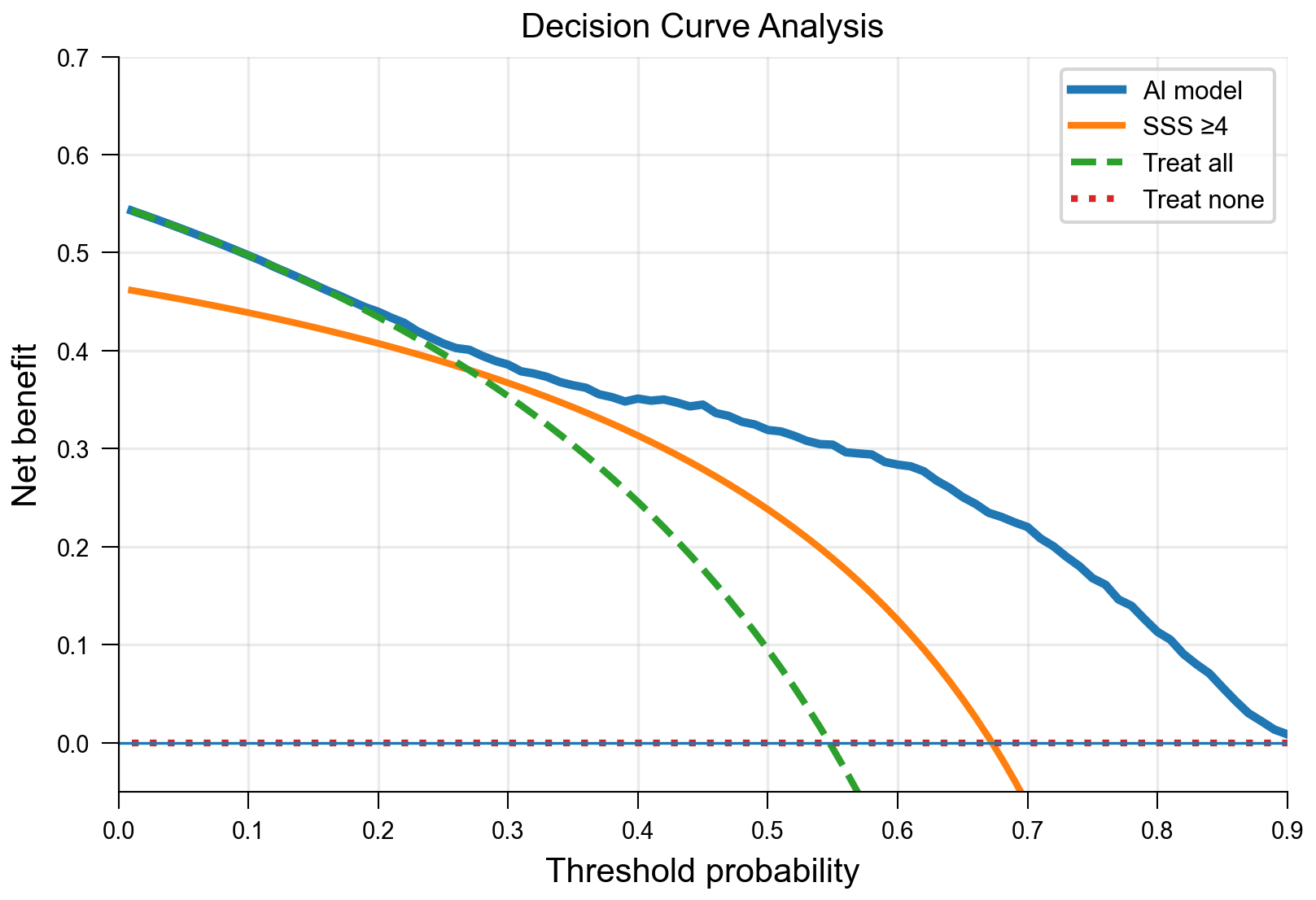

**Supplementary Figure 2: Decision curve analysis.** Net benefit of the AI model compared with SSS ≥4 and treat-all/treat-none strategies across threshold probabilities in the external diagnostic validation cohort. For SSS, net benefit was calculated using the fixed conventional abnormality threshold of SSS ≥4 across all threshold probabilities. SSS, summed stress score.

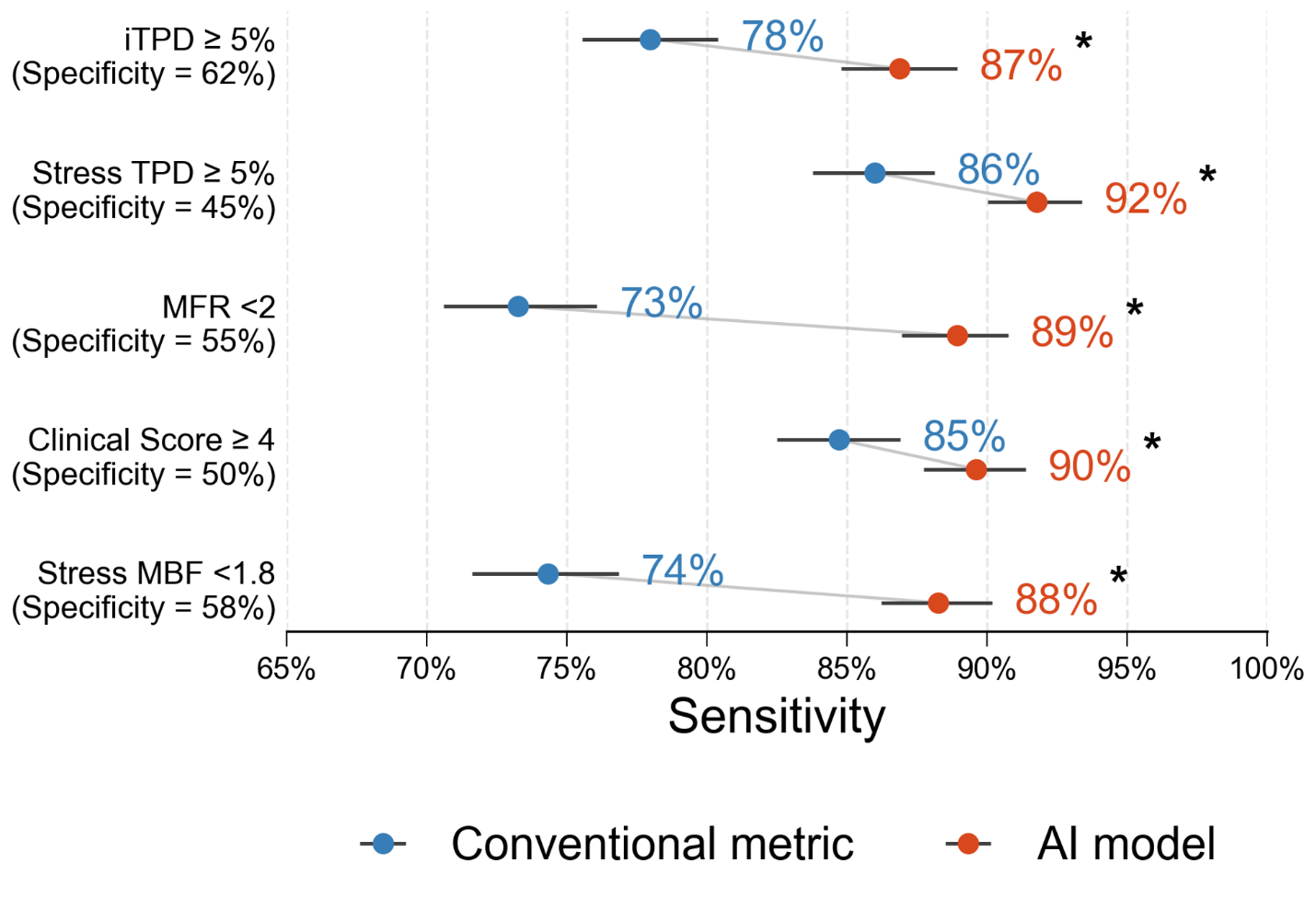

**Supplementary Figure 3: Sensitivity of the AI model at specificity matched to individual conventional PET metrics.** For each comparison, a separate AI-CAD operating threshold was selected to match the specificity of the corresponding conventional metric. Points indicate sensitivity estimates, error bars indicate 95% confidence intervals, and connecting lines show paired comparisons. MFR and MBF were selected from the minimum of the 3 vessels. Asterisks indicate significantly different sensitivities by McNemar’s test (P < 0.05). AI, artificial intelligence; iTPD, ischemic total perfusion deficit; MBF, myocardial blood flow; MFR, myocardial flow reserve; SSS, summed stress score; TPD, total perfusion deficit.

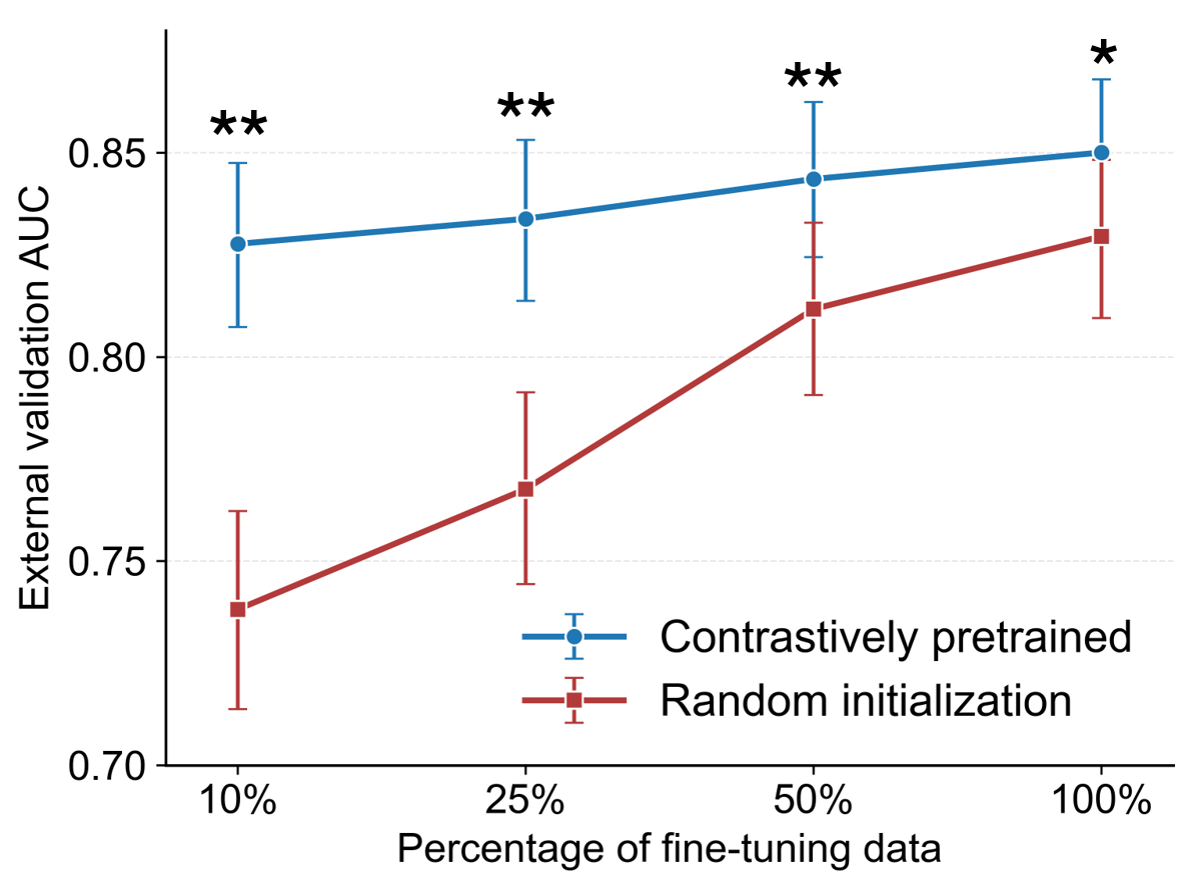

**Supplementary Figure 4: Label efficiency of contrastive pretraining.** External validation AUCs of contrastively pretrained and randomly initialized models after fine-tuning with 10%, 25%, 50%, or 100% of the angiography-labeled cohort. The same nested patient subsets were used for both models at each fraction. Error bars indicate 95% confidence intervals. Asterisks indicate significantly different AUCs by paired DeLong test (* P < 0.05, ** P< 0.001).
